# scRNA-Seq reveals elevated interferon responses and TNF-α signaling via NFkB in monocytes in children with uncomplicated malaria

**DOI:** 10.1101/2023.06.02.23290878

**Authors:** Collins M. Morang’a, Riley S. Drake, Vincent N. Miao, Nancy K. Nyakoe, Dominic S.Y. Amuzu, Vincent Appiah, Yaw Aniweh, Yaw Bediako, Saikou Y. Bah, Alex K. Shalek, Gordon A. Awandare, Thomas D. Otto, Lucas Amenga–Etego

## Abstract

Malaria causes significant morbidity and mortality worldwide, disproportionately impacting sub-Saharan Africa. Disease phenotypes associated with malarial infection can vary widely, from asymptomatic to life-threatening. To date, prevention efforts, particularly those related to vaccine development, have been hindered by an incomplete understanding of which factors impact host immune responses resulting in these divergent outcomes. Here, we conducted a field study in 224 malaria positive individuals (rapid diagnostic test - RDT) from a high transmission area in Ghana, to determine immunological factors associated with uncomplicated malaria “patients” compared to healthy individuals in the community “controls”. Generally, the patients had higher parasite density levels although it had a negative correlation with age, suggesting that, is a key indicator of disease pathogenesis. We applied single-cell RNA-sequencing to compare the immunological phenotypes of 18,176 peripheral blood mononuclear cells (PBMCs) isolated from a subset of the patients and controls (n=11/224), matched on location, age, sex, and parasite density. On average, patients were characterized by a higher fractional abundance of monocytes and an upregulation of innate immune responses, including those to type I and type II interferons and tumor necrosis factor-alpha (TNF-α) signaling via NFκB. Further, in the patients, we identified more putative interactions between antigen-presenting cells and proliferating CD4 T cells and naïve CD8 T cells driven by MHC-I and MHC-II signaling pathways, respectively. Together, these findings highlight transcriptional differences between immune cell subsets associated with malaria that may help guide the development of improved vaccines and new therapeutic interventions for individuals residing in endemic areas.

## Background

In 2022, global estimates of malaria cases and deaths have increased to 249 million cases and 608 000 deaths ^1^. However, the development of an effective vaccine to address this global health threat remains challenging due to an incomplete understanding of the parasite’s biology and limited knowledge of which host factors influence clinical responses to infection.

In malaria endemic communities, individuals may harbour malaria infections with mild to no symptoms warranting treatment, here referred to as healthy community controls. Such infections may be cleared naturally or progress to a uncomplicated malaria, where symptoms become profound enough to necessitate medical intervention. Instructive factors include environmental exposures, transmission intensity, host and parasite genetics, host-pathogen interactions, and host immune responses ^2–5^. Illustratively, upregulation of interferon responses and p53 gene expression can attenuate inflammation and protect children from fever ^6^; and, when comparing children with asymptomatic and severe malaria, the genes most upregulated in severe cases are related to immunoglobin production and interferon signaling ^7^. As reviewed previously, studies have postulated that interferons can orchestrate immune regulatory networks to dampen inflammatory responses and restrict humoral immunity, thus playing a critical role as a wedge that determines protection versus permissiveness to malaria infection ^8,9^.

Similarly, it has been shown that the number and phenotype of cells responding to infection can vary with exposure to *Plasmodium* ^10^. For example, Africans, who tend to have higher levels of exposure, have been shown to exhibit metabolic and platelet activation during malaria infection as compared to typically infection-naïve Europeans ^10^. Similarly, children who experience high cumulative malaria episodes show upregulation of interferon-inducible genes and immunoregulatory cytokines, suggesting an immune modification to prevent immunopathology and severe outcomes during new infections ^11^. Beyond differences in exposure and infection history, the strain responsible for each infection can also alter immune response dynamics and disease pathogenesis ^12,13^.

Since so many factors can influence host response dynamics to infection (e.g., exposure, the timing of infection), some studies have implemented tightly regulated models of malarial infection, such as controlled humanlZlmalaria infections (CHMI). CHMI studies have identified several pathways, including toll-like receptor signaling ^14^, platelet activation ^10^, interferon signaling ^10,15,16^, and B-cell receptor signaling, that are involved in immunological modulation of *Plasmodium falciparum* infections ^6^. Although CHMI enable more controlled examinations of host-pathogen dynamics post-malaria infection, clinically relevant differences can arise between responses seen in CHMI and natural exposure due to unresolved immunopathological mechanisms elicited during *P. falciparum* infection ^10^. Thus, studies involving natural cohorts provide a better avenue to understand vaiability in immune responses developed through repeated exposure and how they influence disease phenotypes. Besides, immunity to malaria develops very slowly through repeated infections, and can wane quickly if individuals leave a malaria endemic areas, suggesting that continuous natural exposure to malaria antigens is important for development of long term immunity ^11^. Collectively, these studies demonstrate the importance of obtaining a more comprehensive understanding of the host and pathogen factors that influence immune responses to inform the development of new therapeutic approaches and improved vaccines.

To date, most genomic analyses of immunological responses to malarial infection have been performed in heterogeneous cell populations of blood, brain, liver, or spleen tissues ^17^. The majority of these studies have been conducted in children and the studies show that symptomatic infections, as mentioned above, are characterized by upregulated expression of genes involved in interferon signaling, antigen presentation, neutrophil-associated signatures, and B cell modules relative to healthy controls ^17^. Adults, meanwhile, present slightly varying responses: symptomatic Malian adults, compared to naïve individuals, had upregulated B cell receptor signaling but more modest upregulation of interferon responses, while symptomatic Cameroonian adults showed marked induction of genes related to interleukins and apoptosis compared to presymptomatic individuals ^18,19^. These inconsistencies may be related to patient history/exposure or differences in cellular composition influencing clinical course through a combination of direct and indirect responses. The emergence of single-cell transcriptomics provides a unique opportunity to examine the sources of this variability ^20^ by profiling abundance and transcriptomic variation across immune cell populations in individuals with high malaria exposure but divergent clinical phenotypes. Moreover, by examining the expression of ligands, receptors, and genes involved in intercellular signaling, we can identify the critical mediators of immune responses and the pathogenesis of malaria for subsequent validation ^21^.

Here, we present a comparative analysis of peripheral blood mononuclear cells (PBMCs) phenotypes in children from two related surveys conducted in 2019. An active case detection of *P. falciparum* infections at the community level (controls) and passive case detection at the health facility level for patients with uncomplicated malaria (patients) in an endemic area in northern Ghana. Our data describe in unprecedented detail, cell subsets and signaling pathways associated with disease severity to provide new insights into the immune response mechanisms that influence the course of *P. falciparum* infections in young children.

## Results

### Clinical characteristics of study participants

In this study, we defined “controls” as healthy individuals who tested *Plasmodium* positive by RDT in the community. We defined “patients” as individuals with uncomplicated malaria who visited the hospital/health center from the same community and tested *Plasmodium* positive by RDT and were treated on outpatient basis. All samples were collected from the same region, Upper East region of Ghana which is a high transmission area. Overall, 224 individuals were surveyed, including 40% (90/224) of the participants who were community healthy controls and 60% (134/224) of the participants who were patients with uncomplicated malaria **(Figure 1a, Supplementary Table 1).** Although most participants were children between 1-15 years, there was no significant difference between the median age of patients compared to the controls (Wilcoxon rank-sum test, P=0.74) ***(Supplementary Table 1)***. But, there was a significant difference in the median parasite density of patients compared to the controls (Wilcoxon rank-sum test, P<0.001) ***(Supplementary Table 1)***. Further, the study sought to determine if the patients had higher parasite densities than the controls regardless of age. In general, there was negative correlation between parasite density and age regardless of phenotype up to age 25 years **(*Figure 1b*).** Under 3 years, the patients tended to have lower parasite densities, but these were still higher than their controls counterparts. After about age 10 parasite densities fell gradually and plateaued around age 25 but with high variability between the groups **(*Figure 1b*)**. The correlation between parasite density and malaria patients was statistically significant (R2 = −0.38, P<0.001), but the correlation between parasite density and age was not statistically significant in the control group **(Figure 1b)**.

**Figure 1:**
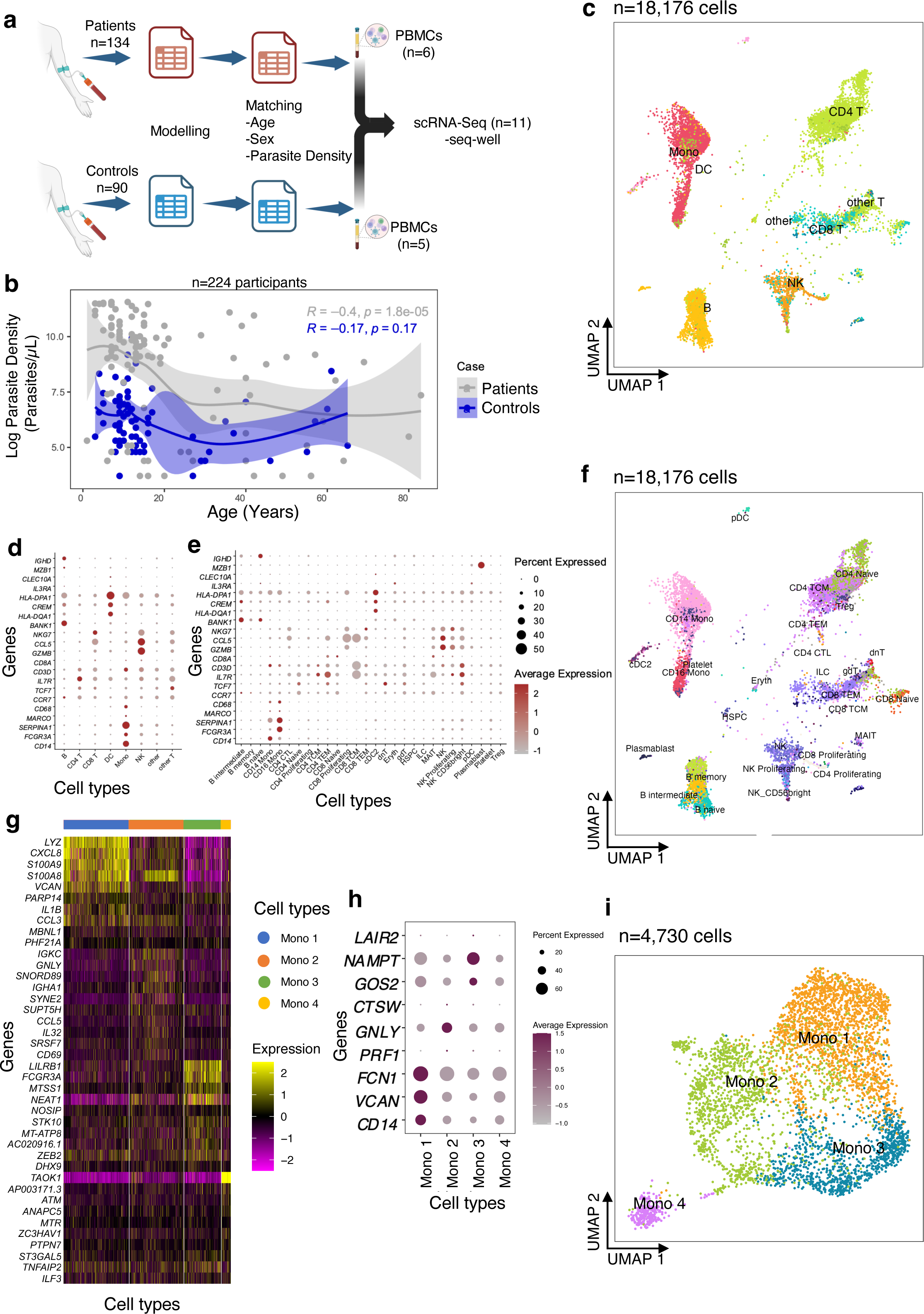
Analysis of scRNA-Seq data from uncomplicated malaria patients and community healthy controls. **a)** Experimental flow showing that PBMCs were collected from eleven individuals out of 224 based on the modelling and matching the patients and controls by age, sex, and parasite density. **b)** Regression analysis between parasite density and age for patients (grey) and controls (blue). **c)** Uniform manifold approximation and projection (UMAP) plot of 22,819 cells from eleven participants colored by identities of 10 cell clusters; mainly B cells, T cells, and Mono. **d)** Expression levels of cluster-defining marker genes organized by color intensity to show the average expression of the marker in that particular cell type and the proportion of cells with non-zero expression shown by the size of the dot. **e)** Markers used to annotate the subclusters to various cell subsets showing average expression and fraction of cells expressing the marker. **f)** Reference mapped dataset showing the predicted subclusters of B, CD4 T, CD8 T, NK, Mono, and DC cell subsets. Reference-defined cell subsets were generated from CITE-seq reference of 162,000 PBMCS measured using 228 antibodies ^45^. **g)** UMAP of re-clustered and re-embedded Mono showing four subclusters of the CD14 and CD16 Mono. **h)** Markers used to identify monocyte subclusters. **i)** Mono top 10 highly expressed genes in each subcluster.

### Profiling pediatric malaria immune-cell populations using single-cell analysis

In order to examine global differences in cellular composition, gene expression and intercellular communication between the two groups, we matched individuals based on age (aged 4-8 years), sex, and parasite density for both patients and controls and performed single-cell RNA-seq (scRNA-seq) (***Figure 1a, Supplementary Table 2)***. There was no significant difference in median parasite density between patients and controls in the matched individuals (Wilcoxon rank-sum test, P>0.71) (***Supplementary Table 2)***. In total, we generated 18,176 high-quality single-cell profiles across eleven children with *P. falciparum* infections, allowing us to ascertain differences in expression patterns of immune response genes that might influence disease pathogenesis. Each sample was profiled using Seq-Well S^3^, a portable, simple massively parallel scRNA-Seq method ^22^. The resulting data were filtered to remove cells based on the fractional abundance of mitochondrial genes (<30%) and transcripts expressing in <20 cells. After variable gene selection, dimensionality reduction, clustering, cluster removal, and reclustering (***Methods***), we retained 18,303 transcripts and identified 10 distinct cell subsets in the 18,176 cells, across the two groups of children (***Figure 1c; Supplementary Figure 1a***).

We manually annotated these 10 clusters using known RNA marker genes to identify B cells, CD4 T cells, CD8 T cells, natural killer (NK) cells, monocytes (Mono), and dendritic cells (DC) (***Supplementary Figure 1b and 1c***). To identify and enumerate cell subsets in our dataset at higher resolution, we opted to map our query dataset to an annotated multimodal reference dataset of PBMCs. First, we confirmed that all the cell subsets identified using manual annotation were present in the resultant UMAP (***Supplementary Figure 1d***). As expected, our reference mapped dataset recapitulated all PBMC subsets, including B, CD4 T cells, CD8 T cells, NK cells, Mono, and DC (these subsets are used throughout the work; ***Figures 1c and 1d***). We identified several subclusters, such as intermediate, memory, and naïve B cells; naïve, proliferating, effector memory and central memory CD8 and CD4 T cells; proliferating CD56+ NK cells; CD14+ and CD16+ monocytes (Mono); plasmacytoid (pDC) and conventional (cDC) dendritic cells, and other cell subsets (***Figures 1e and 1f***). Since the reference dataset has only annotated two Mono clusters (CD14+ and CD16+), we hypothesized that there might be additional transcriptional heterogeneity describing actively responding Mono subpopulations. Therefore, further sub-clustering was done which resolved the Mono into 3 large subpopulations (Mono 1, Mono 2, Mono 3) and 1 small cluster (Mono 4) (***Figures 1g, 1h and 1i***) based on previously reported markers ^23^. Taken together, these data distinguish nearly all distinct cell subsets that were present in PBMCs of children in both the patients and controls.

### Differences in relative cellular composition between the groups

Next, we asked whether there were significant differences in the relative proportions of cell types between the patients and control group. We found that relative cell proportions of the major cell subsets (B, CD4 T, CD8 T, NK, Mono, and DC) varied between individuals in each group (***Figures 2a and 2b, Supplementary Table 3***). The patients exhibited elevated levels of circulating Mono while the controls had higher proportions of circulating B cells (Dirichlet-multinomial regression, *P*<0.01; ***Figure 2a, Supplementary Table 3***). Further analysis of the B cell subsets showed that the abundance of naïve and intermediate B cells was significantly reduced in the patient group compared to the control group (Dirichlet-multinomial regression, *P*<0.05; ***Figure 2b, Supplementary Table 3***). We also found a significant expansion of both CD14+ and CD16+ Mono subsets in patients compared to the control group (Dirichlet-multinomial regression, *P*<0.01; ***Figure 2b, Supplementary Table 3***). Although there is evident variation in cellular proportions of T lymphocytes among all the individuals (***Figures 2a and 2b***), we did not observe any significant difference in proportions of either CD4 or CD8 T cells between the groups (Dirichlet-multinomial regression, *P*>0.05; ***Figure 2b, Supplementary Table 3***). However, the proportions of naïve and central memory CD4 T cells were significantly higher in the patients compared to the control group (Dirichlet-multinomial regression, *P*<0.01; ***Figure 2b, Supplementary Table 3***). NK cell frequency was also higher in patients suggesting that they may play a role in disease progression (Dirichlet-multinomial regression, *P*>0.05; ***Figure 2b, Supplementary Table 3***). Among NK cells, the proliferating and CD56+ subsets were higher in patients compared to controls, but these differences were not statistically significant (Dirichlet-multinomial regression, *P*>0.05; ***Figure 2b, Supplementary Table 3***). Overall, the minor subsets of T cells and other cell types with low frequencies did not show differences in proportions between the groups but the main cell subsets had significant differences in proportions between patients and controls.

**Figure 2:**
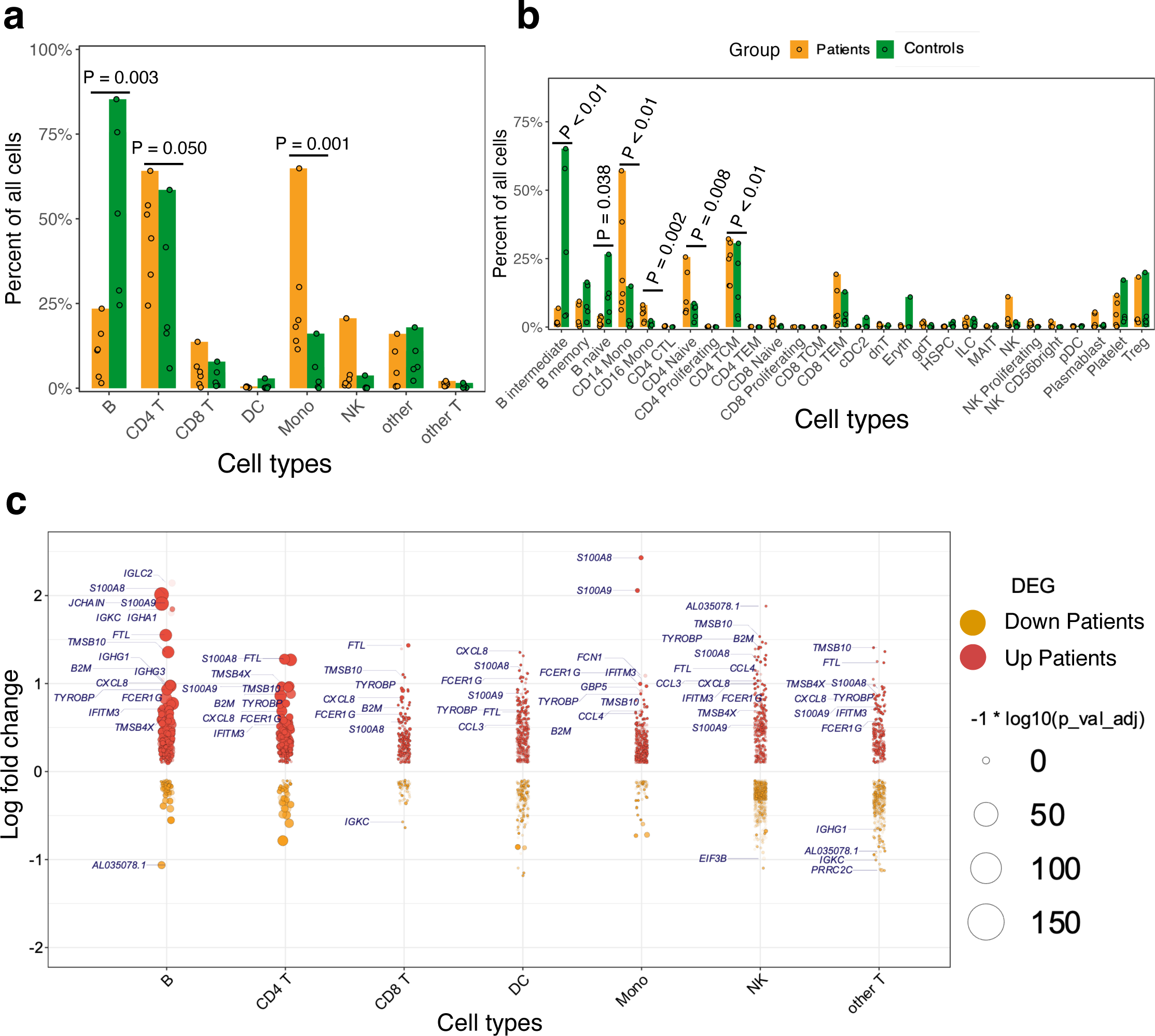
Profiling of immune cells from patients compared to the controls. **a)**. Relative cell proportions of the major cell subsets within patients and control groups. Statistical tests were conducted using the Dirichlet Multinomial Regression in the DirichletReg package in R ^47^. The dots represent individual proportions while the color scheme represents the patients and control groups. **b)** Relative proportions of minor cell subsets compared between patients and controls. Cell proportions per group and P-value are shown in **Supplementary Table 1. c)** Violin-like plots showing genes that are differentially expressed between patients and controls. The x-axis shows the Log_2_ fold change against the cell subsets (y-axis) – i.e., B cells, Mono, CD4 T cells, CD8 T cells, other T cells, dendritic cells (DC) and natural killer (NK) cells. The color scheme is based on the upregulated (up patients) and downregulated (down patients) genes in patients and the size of the point represents the adjusted P value. The frequency shows the number of comparisons in which the gene is significantly expressed in the cell subset.

### Comparative analysis of inflammatory responses in children with malaria

Having identified shifts in the composition of circulating immune cells between the patients and controls, we next asked whether gene expression differed within each immune cell subset between the two groups. Comparing patients to controls, we observed the largest transcriptional changes (measured by pairwise DE across cell types with adjusted P value < 0.05 and log fold change > 2) within B cells and Mono (***Figures 2c, Supplementary Table 4***). Apart from B cell function genes, there was a general trend towards upregulation of inflammatory genes in B and T cells in patients relative to control group, including *S100A8, CXCL8,* and *S100A9* (***Figure 2c***). Significant transcriptional changes were also observed in Mono, with genes such as *IFITM3, FCER1G,* and *CCL4* being upregulated in patients compared to the control group (***Figure 2c***). Patients were also associated with the upregulation of Major Histocompatibility Complex I (MHC-I) genes such as *HLA-A* and *HLA-C* which are involved in antigen presentation in Mono (***Figure 2c***). In CD4 and CD8 T cells, there was increased expression of some inflammatory factor signalling genes such as *CXCL8* and *NFKBIA* in patients relative to the control group, suggesting direct sensing of parasite products during clinical presentation (***Figure 2c***). Using gene set enrichment analyses (GSEA), we found that the patients had robust induction of several innate immune response pathways such as TNF-α signaling via NF-κB, TGF-β signaling, IL6-JAK-STAT pathway, complement, IL2-STAT5 signaling, inflammatory response, interferon-α response (IFN-α), and interferon-γ response (IFN-γ) (***Figure 3a, Supplementary Table 5***). We observed that although each cell type was enriched in one or more of these pathways, there was a unique molecular signature of the genes involved in each. Upregulation of IFN-γ and IFN-α response pathways in Mono were characterized by increased expression of genes such as *IFITM2, IFITM3, IL10RA,* and *TNFAIP3,* while in NK cells they were typified by genes such as *NFKBIA, CD69*, and *ISG20* (***Figure 3b* *& 3c, Supplementary Table 5***). Mono and natural killer cells upregulated TNF-α signaling via the NF-κB pathway with the induction of genes related to this pathway such as *IL1B* and *TNFAIP3* for Mono, and *IL7R, CD44,* and *NFKBIA* for NK cells (***Figure 3b* *& 3c, Supplementary Table 6***). Inflammatory responses in Mono were characterized by *IL10RA, IL1B*, and *CXCL8* while in NK cells they were driven by *CD69, IL7R, CXCL8,* and *NFKBIA* among others (***Figure 3b* *& 3c, Supplementary Table 5***). Thus, the enrichment of unique genes for each cell subset for similar pathways suggests a specific but concerted contribution of each cell subset toward the innate immune response in patients.

**Figure 3:**
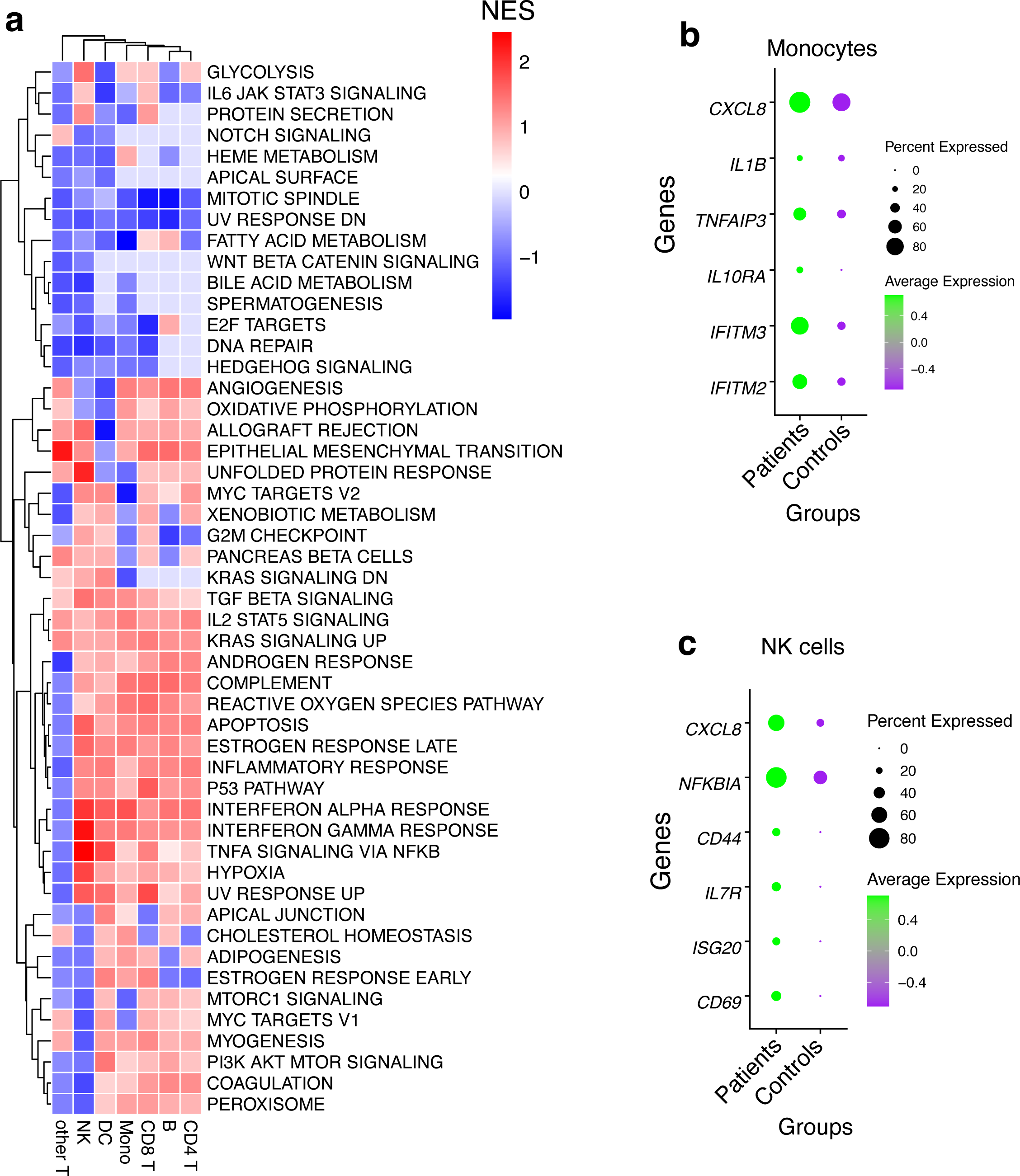
Pathway analysis using geneset enrichment method. **a)** Pathway analysis using an immunologic signature geneset enrichment analysis (GSEA) and the color scheme is based on the normalized enrichment score of genes DEG in patients. **b)** Dot plots showing some of the leading edge genes in IFN-γ and IFN-α response, TNF-α signaling via NFκB & inflammatory response pathways in Mono and, **c**) NK cells. Dot size represents the fraction of cell subsets expressing a given gene. The dot color indicates scaled average expression by gene column.

### Relative enrichment of ISGs gene modules in monocytes of patients relative to controls

Since IFN genes were significantly upregulated in Mono patients relative to the control group, we next sought to determine if entire gene modules were enriched. Interferon stimulated genes (ISGs) modules scores were significantly higher in B cells, DC, CD4 T cells, and Mono in patients compared to the control group (***Wilcoxon, adjusted P < 0.01 for all comparisons, Figure 4a, 4b, 4d, and* *4e***); however, there were no significant differences in ISG module scores in CD8 T cells and NK cells. Further examination of intra- and inter-individual variation in these module scores revealed substantial intra-individual variation in cells from the same participant and between cells of the same type from different participants (***Supplementary* Figure 1e**). Overall, our data show that Mono play a significant role in defining malaria patients compared to control participants from the same community through induction of the ISGs gene modules.

**Figure 4:**
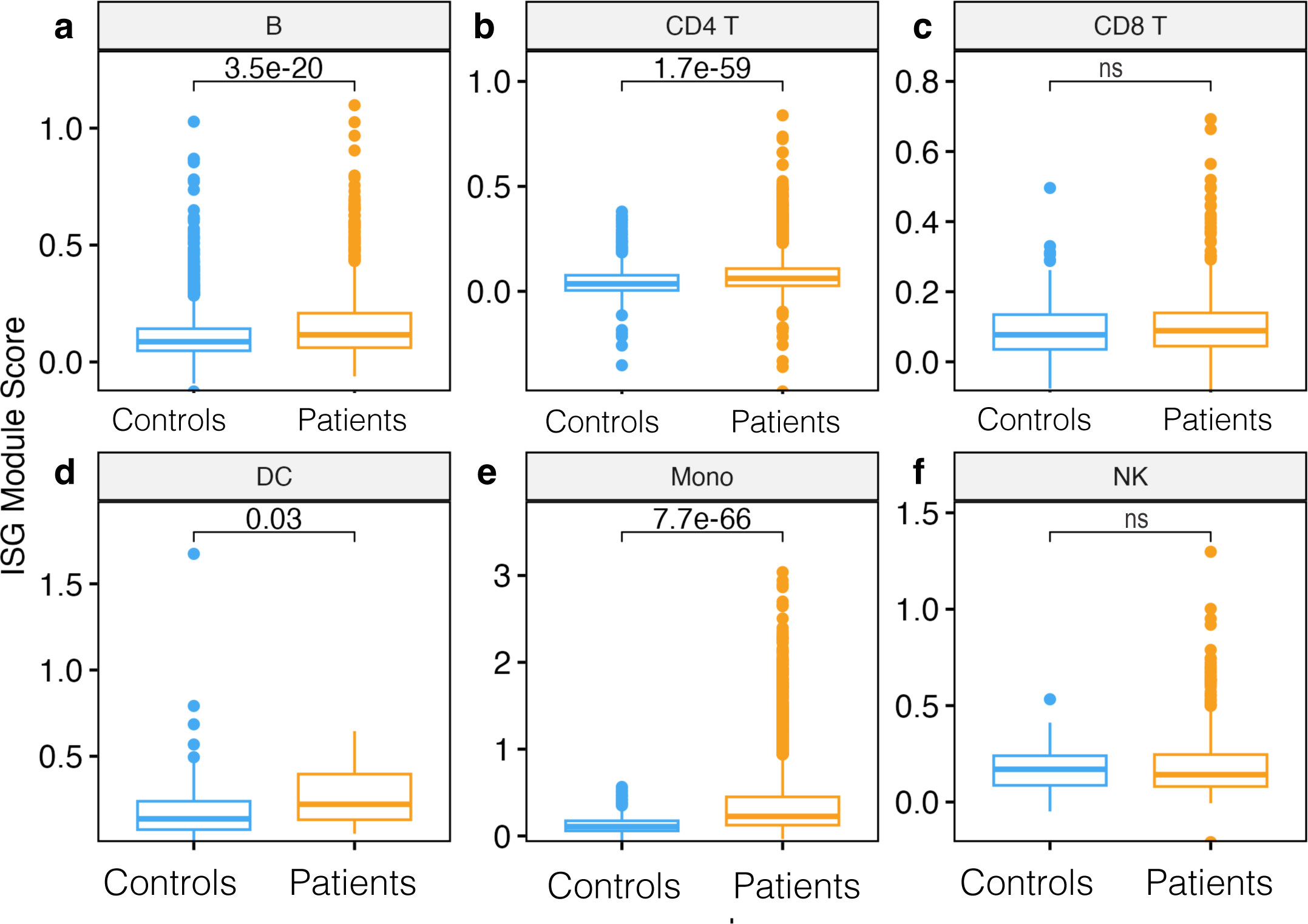
Module score analysis of innate immune gene modules. **a)** Boxplot showing interferon-stimulated gene (ISG) module scores per cell subset compared between patients and controls in B cells, **b)** CD4 T cells, **c)** CD8 T cells, **d)** DC e) Mono, **f)** NK cells. Module scores are computed using the AddModuleScore function in the Seurat R package. Statistical significance between the patients and controls of each cell subset was computed using Wilcoxon sign-rank test with Bonferroni correction. Non significant differences are indicated by ns.

### Role of MHC-I and MHC-II signaling pathways in cell-to-cell interactions

Next, we used our single-cell data to infer putative axes of cell-to-cell communication using signaling ligands, cofactors, and receptors. First, we discerned cell-to-cell interactions in the patients and found that the number of interactions (ligand-receptor) originating from primary innate immune cells such as DC and Mono were greater than those originating from non-antigen presenting cells (***Supplementary Table 6***). However, our data show very few inferred cellular communication networks in the control group (***Supplementary Table 6***). This analysis suggests a role for Mono as antigen-presenting cells in orchestrating pro-inflammatory responses by interacting with proliferating CD4 T cells, intermediate B cells, effector memory T cells, and naïve CD8 T cells in the patient group (***Figure 5a***). Conventional DC also produced factors that interact with proliferating and effector memory CD4 T cells respectively, suggesting a concerted effort by antigen-presenting cells to activate the immune response in patients (***Figure 5a***). Communication probabilities indicated that MHC-I and MHC-II play a role in these interactions among other pathways. The most significant receptor-ligand pairs for HLA-A, HLA-B, HLA-C, HLA-E, HLA-G, and HLA-F ligands for MHC class I include CD8A, CD8B, LILRB2, and LILRB1 (***Figure 5b***). The leading intercellular ligand-receptor pairs with CD4 T cells as signal receivers were distinct HLA genes, with the highest relative contribution being driven by HLA-DRA and HLA-DRB1 (***Figure 5c***). The other minor signaling pathways that were important in patients include MIF, RESISTIN, ANNEXIN, GALECTIN, ADGRE5, APP, CD22, CD45, SELPLG, CD99, CLEC, and TNF signaling networks. For the TNF signaling pathway, the CD56^+^ NK cells showed to be interacting with Mono and also with proliferating CD4 T cells, effector memory CD4 and CD8 T cells, and cDC (***Figure 5d***). This cell communication network was mediated by *TNF* in the sender cells and *TNFRSF1B* in the receiver subsets (***Figure 5e***), and this corroborates the DE results (***Figure 5g***). We examined the expression levels of *TNFRSF1B* across all the cell subsets and found that indeed it was expressed in all the receiver cells (***Figure 5f***). Only the pDC and CD16 Mono showed cell-to-cell interactions with naïve and intermediate B cells and might be playing a role in B cell activation and development in the control group through MHC class II molecules (***Figure 5g and 5h***). Therefore, exposure of innate immune cells to parasite ligands may potentially activate intracellular signaling cascades through cell-to-cell interactions to induce rapid expression of a variety of innate immune genes.

**Figure 5:**
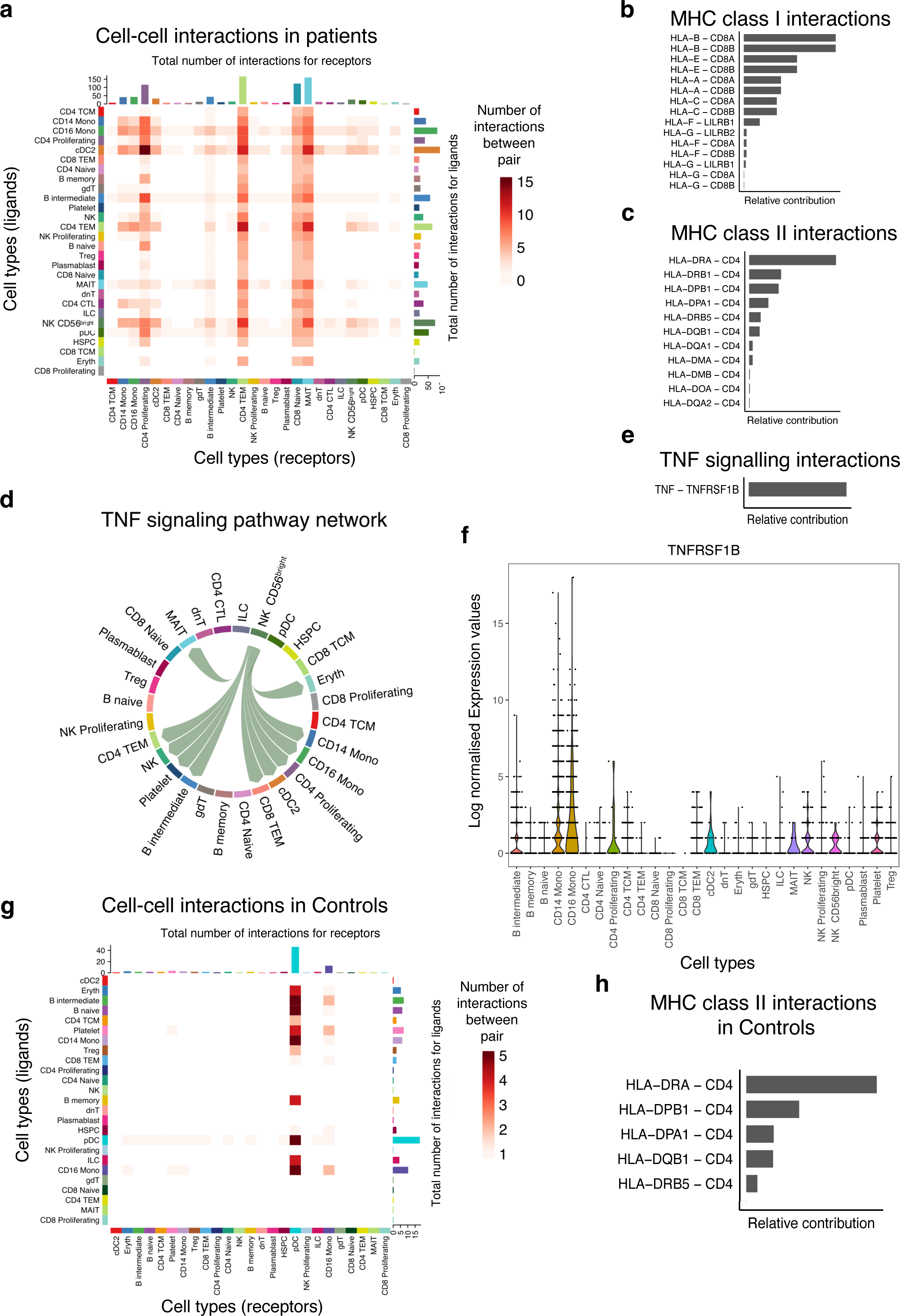
Primary innate immune cells dominate the cell-to-cell interactions with other cell subsets. **a)** Heatmap showing the number of interactions between the PBMCs cell subsets. The y-axis shows the signal senders and the x-axis shows the signal receivers. **b)** Relative contribution of ligand receptor pairs in patients within the MHC class I signaling pathway and **(c)** MHC class II signaling networks, respectively. A higher relative contribution indicates the magnitidue of contribution of the ligand-receptor and its significant role in the MHC I or II signalling networks. **d)** Cell communications through the TNF signaling pathway and the arrows indicate signal sender to receiver. **e)** Relative contribution of the TNF-TNFRSF1B ligand-receptor pair towards the TNF signaling pathway. **f)** Violin plots showing the expression levels of the TNFRSF1B in the Seurat object for the cell subclusters. **g)** Heatmap comparison showing the overall signaling between all cell subclusters and the number of interactions. **h)** Relative contribution of MHC class II signaling pathway in the control group.

## Discussion

Here, we recruited 224 participants malaria from Navrongo, a high malaria transmission area with seasonal fluctuations ^24^. Interestingly, most of participants indicated that they use long lasting insecticide treated mosquito nets (LLINs), which helps to explain the low frequency of infections; after screening 1,000 individuals in the community, <10% of them were positive for *P. falciparum* as community healthy controls, suggesting a reduction in the malaria infection reservoir. The National malaria elimination programme (NMEP) distributes LLINs as part of strategy interventions, including community-based seasonal malaria chemoprevention initiatives for children under 5 years in order to reduce the malaria burden in this area ^24^. The ability of insecticide treated nets (ITNs) to interrupt malaria transmission has been shown in large scale studies, which demonstrated that modern housing and ITNs could reduce malaria infections by 1% and 16%, respectively ^25^. Further, we investigated the relationship between age and parasite density, and found that parasite densities tended to decrease with age, but the levels were generally higher in patients compared to controls in this high transmission intensity area ^5^.

To better understand cellular responses driving these divergent clinical phenotypes, we performed scRNA-seq on PBMCs samples from eleven of the 224 individuals among the two groups, controlling for group variability driven by age, fever and parasite density. This enabled us to identify a potential role for interferon responses and TNF-α signaling via NFκB in Mono during the clinical manifestation of pediatric malaria infection. We also found differences in the fractional abundances of PBMC cell subsets, with patients characterized by a proportional increase in Mono while controls had a higher proportion of circulating B cells. We showed cellular level variations in the expression of innate immune modules within and between individuals as well as between clinical phenotypes. Further, we identified a role for Mono and other innate immune cells through MHC-I and MHC-II molecules in driving cell-to-cell interactions with CD8 and CD4 T cells respectively. Together, our work recontextualizes the function of the innate immune cells in malaria, demonstrates how variable their responses can be, and links specific acute phase response signaling pathways to clinical presentation.

Differential gene expression comparing patients and controls across cell types revealed a significant upregulation of genes associated with innate immunity in different cell types. We show that CCL3 and CCL4 (also known as macrophage inflammatory protein MIP-1α and MIP-1β respectively) were upregulated in Mono of patients, suggesting their possible role in modulating clinical disease ^26^. CXCL8, the most potent human neutrophil attracting/activation chemokine ^27^ was also highly upregulated in B cells, CD4, and CD8 T cells. Other studies have shown that circulating levels of CXCL8 and CCL4 correlated with parasite density, and when found in the cerebrospinal fluid they can predict cerebral malaria mortality ^13,28–30^. Furthermore, the adaptive immune cell subsets (B cells and T cells) in the patient group expressed two alarmins (S100A8 and S100A9) that are known to form calprotein heterodimer, an endogenous TLR4 ligand; this could suggest a possible role to silence hyperinflammation ^31^. We also show significant expression of FCER1G in B cells, Mono, and DC in patients, which is induced by IFN-γ and encodes for a gamma chain of the FC receptor and it is suggested to play an important role in controlling parasitemia ^6^. Collectively, our data imply that both adaptive and innate immune cells cooperatively play a role during the pathogenesis of malaria in patients when compared to healthy controls.

We showed that several immune-related pathways are activated by *Plasmodium* infection and disease including the TNF-α signaling via NFκB pathway, IFN-α/γ responses, IL2-STAT5 signaling, and inflammatory response pathway in patients. Since the parasite life cycle involves repeated red cell invasion and rupture, the release of pyrogenic cytokines that drive these pathways such as interleukins, interferons, and TNF in Mono and NK cells, can signify pathophysiological events occurring in malaria patients ^13,32^. These observations could also mean that children who patients were sampled quite early during the onset of the disease progression trajectory ^12^. Our data are consistent with those previously described by integrating whole blood transcriptomics, flow cytometry, and plasma cytokine analysis ^6^, and our results further identify the cell subsets in which these pathways were more enriched. We show that each of the cell subsets has a unique signature of genes enriched in these immunogenic pathways with minimal sharing. Several studies have shown similar innate immune response pathways in individuals with malaria such as whole blood transcriptomics of the Fulani of West Africa ^33^, children repeatedly exposed to malaria ^6,11^, controlled human malaria infection (CHMI) studies ^16^, and even mice models ^31^. We have now confirmed some of these observations and demonstrated that in the patient state, robust upregulation of certain genes in specific cell subsets is associated with systemic inflammatory responses. Innate immune cells, such as Mono, DC, and NK cells, appear to be most reactive in patients, probably due to continuous exposure in a high transmission area as suggested by other studies ^7,31,34^.

By collating gene modules of interferon-stimulated genes (ISGs), we show that there is a differencial expression between patients and controls across different cell subsets. ISGs are normally produced as a function of interferon responses (IFNs) ^8^, which we observe to be enriched in patients. IFNs are produced primarily by DC to activate ISGs in other cells ^35^, and we observed that B cells, T cells, Mono, and DC have higher ISG module scores in patients compared to controls. Notably, our data show that each cell or cell subset responds differently upon IFN activation with varying transcriptional responses of an ISG module between individuals. This variability was also observed for cytokine modules, NF-κB target modules, and HLA modules. Similarly, a previous CHMI study observed striking inter-individual variation in immune cell composition and immune responses, demonstrating that an individual can have a unique immune fingerprint ^10^. Thus, the variations in immune responses that we observed could be attributed to the complexity of the *P. falciparum* life cycle with several developmental erythrocytic stages, duration of infections, intensity of infection in each individual, genetic factors, genetic variation in immune response genes among other factors ^12,36^. These findings on inter-individual variability in immune responses could provide insights when considering the design and evaluation of interventions that target host immunity in the control of malaria.

Our scRNA-Seq data enabled us to quantitatively infer and analyze cell-to-cell communication networks across all the innate and adaptive immune cells (Jin et al., 2021). This analysis enabled us to uncover coordinated interactions between innate and adaptive immune cells through various ligands. The cell-to-cell interactions in patients were driven by MHC class I and II signaling pathways, whereby antigen-presenting cells were shown to have more interactions with proliferating CD4 and naive CD8 T cells. The importance of HLA genes has long been demonstrated by Hill and colleagues who associated HLA-Bw53 antigen and DRB1*1302–DQB1*0501 haplotype to independently protect against severe malaria in West Africa ^37^. Thus, our observations on cell-cell interaction involving HLA molecules and T cells support the importance of these molecules during *P. falciparum* infection and disease progression, consistent with the observed varying degrees of interactions in patients compared to control groups. We also showed that within patient group, there are contrasting interactions between various HLA I and HLA II molecules with CD8 or CD4 T cell receptors respectively, which could be related to their tight regulation and antigen-presenting ability ^38,39^. Activation of CD4 and CD8 T cells has been correlated with protective immunity to malaria, and they can differentiate into several functionally distinct subsets in the presence of various cytokines ^40^. It was not surprising that we identified different fractional abundances of CD4 and CD8 T cell subsets in patients compared to control group of children, but we demonstrate that ultimately this results in varying degrees of interactions with Mono or DC. Future work should seek to identify the mechanisms that result in these variations and their impact in orchestrating phagocytic and humoral responses as this critical knowledge gap will be important in developing T cell-based malaria vaccines.

Overall, by using scRNA-seq on PBMCs obtained from patients and controls in a high transmission area, this work sheds light on the interplay between peripheral immune cells during uncomplicated malaria, uncovering the genes and immune pathways in specific cell types that might play a significant role in defining the outcomes of infection. Data presented here demonstrate that the patients with uncomplicated malaria were characterized by the presence of inflammatory response signatures in specific cell types compared to the control group. The results could also suggest that in the control group, a muted innate immune response or disease tolerance mechanism plays a role in enabling children to harbour malaria parasites in high malaria transmission areas without developing uncomplicated malaria ^41^. The findings are relevant for guiding the development of malaria vaccines, as well as immunotherapeutics for alleviating uncomplicated malaria disease and preventing progression to severe disease.

## Methods

### Study design and sample collection

In 2019, we conducted a cross-sectional active case detection survey in the Kassena-Nankana Municipality of the Upper East Region of Ghana, to recruit children with uncomplicated malaria and controls with *P. falciparum* infections. About 1000 community members were screened for malaria infection using a CareStart^TM^ PfHRP2-based malaria Rapid Diagnostic Test (RDT, Access Bio, NY, USA). Positive cases in the community were defined as “controls”, since these individuals hadn’t sought treatment within the past two weeks. Similarly, a passive case detection of uncomplicated malaria cases was carried out using the same mRDTs to screen individuals presenting at the Navrongo War Memorial Hospital outpatient department. Individuals who tested positive for malaria and who provided written informed consent were recruited into the study as defined as “patients”. Five milliliters of whole blood was collected for PBMC isolation and for thick and thin blood smears for parasite identification and quantification using microscopy. Linear regression models was used to determine the relatisohsip between parasite density and age in patients and controls in R (version 4.2.1).

Of the 224 individuals recruited in both arms of the study, five control participants and six patients were selected for single-cell transcriptomic analysis. Due to the observed clear differences in clinical presentation between the groups driven by fever, headache and parasite density, the 11 children selected for single-cell analysis had close similarity in these factors. PBMCs were isolated in ACD tubes and spun at 2,000 revolutions per minute (rpm) for 10 minutes, and the leukocytes layer was transferred to 15 mL. The leukocytes were mixed with phosphate-buffered saline (PBS) and layered on 3 mL of lymphoprep in a 15 mL falcon tube. The layered cells were spun for 30 minutes at 800 g without breaks and harvested carefully by taking the buffy layer into another falcon tube. The PBMCs were washed twice with PBS and stored in freezing media. During thawing, complete media (RP10) with 20% Fetal Bovine Serum (FBS) was prepared by diluting 20 mLs of FBS in 80 mL of Roswellpark Memorial Institute (RPMI) media ^42^. PBMCs were removed from Liquid Nitrogen to the −80 °C freezer and then thawed during each experiment. Thawing was done by placing the vial in a clean water bath at 37°C until a small crystal of frozen cells was visible. The tubes were cleaned with 70% ethanol, and the contents were transferred to 10 mL of RP10 gently to minimise stressing the cells. The cells were centrifuged at 500g for 10 minutes and resuspended in RP10. Cell viability was estimated using Haemocytometer and PBMCs were used after 1 hr of resting in the incubator at 37 °C.

### Seq-Well scRNA-Seq Workflow

Seq-Well scRNA-Seq S^3^ workflow was performed according to the published methods ^22,43^. In brief, 5×10^5^ PBMCs from each patient were dispensed into a single array containing barcoded mRNA capture beads (Supplementary Figure 1). The arrays were sealed with a Polycarbonate Track Etch (PCTE) membrane (pore size of 0.01*µ*M), allowing cells to remain separated through the lysis and hybridization steps. mRNA transcripts were hybridized and recovered for reverse transcription using the Maxima H Minus Reverse Transcriptase in the first strand synthesis step. Exonuclease (I) was used to remove excess primers and mRNA was captured via poly-T priming of the poly-A mRNA. The captured mRNA underwent first-strand synthesis to generate single-stranded cDNA while bound to the beads. Enzymes with terminal transferase were used to create 3’ overhangs and three cytosines. The overhangs are used in template switching, whereby a SMART sequence is appended to the overhang on both ends of the cDNA molecule during the first strand synthesis. Some templates fail to switch, resulting in loss of the mRNA; hence they are chemically denatured using 0.1M NaOH with random octamer with the SMART sequence in 5’ orientation, and a second strand is synthesized. Whole transcriptome amplification of the cDNA was performed using the KAPA HiFi PCR master mix (Kapa Biosystems). Libraries were pooled and purified using AgenCourt AMPure XP Beads. The quality of the library was assessed using Agilent Tape Station with D5000 High Sensitivity tapes and reagents. Samples were barcoded as described in the Nextera XT DNA (Illumina, USA) segmentation method. Tagmentation was important because, after cDNA amplification and clean-up, there are usually very long cDNA molecules that need to be fragmented to be sequenced by Illumina. The Nextera XT DNA tagmentation method is effective and allows for the addition of adaptors and multiplex indexes at both ends of each fragment ^22^. Finally, the amplified library was purified using SPRI beads, pooled, and sequenced using the NextSeq500 kit (Illumina, USA). Paired-end sequencing was performed with a read structure of 20 bp read one, 50 bp read two, and 8 bp index one as recommended for Seq-Well. The targeted sequencing depth was 100 million reads for all samples.

### Processing sequencing reads

The raw data were converted to demultiplexed FastQ files using bcl2fastq (Version 5, Terra Workspace) using the Nextera XT indices and then aligned to the hg19 human genome using STAR aligner (Version 2.7.9) within the Broad Institute DropSeq workflow (Version 11, Terra Workspace). The data was cleaned using Cell Bender (V 0.2.0) with default settings, to remove ambient RNA ^44^. The raw expression matrices and sample information were loaded into the open-source statistical software R (R version 4.2.1). An array with 45,691 gene features for 22,819 cells described data collected across 11 samples. The data were filtered to include only features expressed in more than 20 cells, and the resultant matrix described 18,303 gene features across 22,819 cells. A Seurat (Version 4.0) object was created, and the metadata was added to it to identify the participants ^45^. Cell cycle scoring was performed and computation of the percentage of mitochondria genes before integration. The object from each participant was transformed individually within the object using SCTransform followed by the selection of integration features, finding the anchors, and finally combined integration. Principal component analysis was performed to reduce the dimensionality of the data in order to identify clusters of cells with similar transcriptomic profiles. Clusters and cluster resolution were determined using FindNeighbors and a customized FindClusters function that showed that the best resolution was 0.523, with an average silhouette score of: 0.2 and 11 clusters. One cluster showed no cluster-specific genes and was removed as multiplets, leaving 18,176 cells. The remaining clusters were reclustered and re-embedded, resulting in 10 clusters with a resolution of 0.292, and an average silhouette score of: 0.301. The average number of transcripts and expressed genes were evaluated per cluster using half violin and boxplots. The clusters were projected to a two-dimensional space using the Uniform Manifold Approximation and Projection (UMAP) ^46^ algorithm in Seurat.

### Reference-based mapping

Immune cell subsets were identified using common cell markers to identify the Mono, T cells, B cells, NK cells, DC, and other immune cell populations. Uniform Manifold Approximation and Projection for Dimensional Reduction (UMAP) was used to embed the cell populations and color code based on the expression of surface markers. The clustered PBMC dataset in this study (query) was mapped to a reference CITE-Seq dataset of 162,000 PBMCS measured with 228 antibodies ^45^. The query data were projected into the same dimensional space as the reference dataset, thus separating the cells into the cell types present in the reference dataset. The method first projected the reference data transformation onto the query data, followed by the application of KNN-based identification of mutual nearest neighbors (anchors) between the reference and query. On an L2-normalized dimensional space, the reference data transferred continuous data onto the query data to annotate the scRNA data based on a weighted vote classifier. For visualization, reference-based UMAP embedding was used, considering that all the immune cell populations are well represented.

### Analyzing differences in samples

Cluster/sample composition was calculated to determine the proportion of cells per cluster and per cell type. Cell subsets that were significantly different between patient and control groups were identified by computing Dirichlet Regression using the DirichReg function in DirichletReg Package in R ^47^. Differentially expressed (DE) genes were computed using the FindMarkers function on Seurat (Version 4.0), which we used to determine differentially expressed genes in the patient and control groups using MAST with significance at *(P<*0.05) and log fold change of > 0.2. Control 4 was not included in the DE analysis due to different levels of cytokine module scores compared to the other controls participants (***Supplementary Figure 1e***). DE genes were visualized using volcano-like plots and heatmaps to compare all the cell types between patients and controls. The fgsea (R-package) was used to analyze the pre-ranked gene set enrichment analysis (GSEA). Module scores for HLA genes, ISG, NFκB target genes, and cytokines were analyzed using the AddModuleScore function in the Seurat R package. Statistical differences in module scores between the patients and control groups for each cell subset was computed using Wilcoxon sign-rank test with Bonferroni correction. Boxplots were used to visualize the module scores for each cell, denoting the median and interquartile range.

### Cell-to-cell Interaction using CellChat

CellChat (Version 1.1.1) was used to quantitatively infer and analyze cell-to-cell communication networks ^21^. Statistically significant intercellular communication between cell groups was identified using permutation tests, and interactions with a significance level of less than 0.05 were considered significant ^21^. Heatmaps were used to visualize each signaling pathway and their cell-cell communications, highlighting the number of interactions, the sources (ligands) of the interactions, and the receivers (receptors) of the interactions. The relative contribution of each ligand-receptor pair to the overall signaling was shown in bar plots. The relative contribution provides a measure of a particular ligand-receptor interaction in a particular cell-cell signaling network. This measure demonstrates the importance or significance of the interaction in mediating cell communication between the cell types and potential functional relationships. It is calculated by comparing the expression levels of different cell receptor and ligand genes between the cell types while accounting for all the possible interaction pairs within a signaling network.

## Data Availability

All data generated or analyzed during this study are included in this published article and its supplementary information files. The scRNA-Seq data are available at https://cellatlas-cxg.mvls.gla.ac.uk/PBMC.Pediatric.Malaria.Ghana.

## Acknowledgments

We acknowledge the study participants for contributing to the study, and the staff of Navrongo Health Research Centre, who provided support for this study. We are grateful to Felix Ansah, Jersley Chirawurah, and Jonas Kengne, for their contributions to the data collection and critical review of the work. All data were stored and analyzed on the University of Ghana’s high-performance computing system (Zuputo).

## Author’s contributions

G.A.A, TDO, LA, YB, and AKS contributed to design and conceptualization; CMM, LA, DA, YA, and NKN contributed to sample collection and processing. VM, RD, and CMM contributed to performing Seq-Well experiments; CMM performed the data analysis and drafted the manuscript. VM, VA, SB, NKN, TDO, LA, and AKS contributed to data analysis and drafting of the manuscript. All authors read and approved the manuscript.

## Ethics approval and consent to participate

Ethical clearance was obtained from the Noguchi Memorial Institute of Medical Research, University of Ghana **(IRB 0000908),** and the Ghana Health Service **(GHS-ERC 008/02/19).** All participants provided written informed consent before inclusion in the study.

## Consent for publication

Not applicable.

## Supplementary Tables

**Supplementary Figure 1 | Cluster identification and annotation. a)** UMAP of 22,819 cells from all participants, showing 10 clusters in the dataset (following iterative Louvian clustering). **b)** Dot plots showing genes used to manually annotate the clusters and show the fraction of cells expressing it and the non-zero expression. Dot size represents the fraction of cell types (rows) expressing a given gene (columns). The dot color indicates scaled average expression by gene column. **c**) UMAP colored by various manually annotated clusters based on the cell markers **d)** UMAP showing cell clusters identified from a reference-mapped dataset but labeled with the manually annotated cluster identities. **e)** Heatmap showing overall module score for each cell, and grouped based on each participant and all the cell types, and overall study groups. The color scheme represents a scale for module scores.

**Supplementary Table 1:** Descriptive statistics of all study participants

**Supplementary Table 2:** Descriptive statistics of matched study participants for scRNA-Seq

**Supplementay Table 3:** Dirichlet regression analysis of proportions of various cell types compared between the two groups

**Supplementary Table 4:** Differentially expressed genes between patients and controls

**Supplementary Table 5:** Pathway analysis of differentially expressed genes between patients and control group

**Supplementary Table 6:** Leading pathways in cell-cell interaction analysis for the patient group

**Supplementary Table 7:** Leading pathways in cell-cell interaction analysis for the control group

