## Supplementary figures and images for "scRNA-Seq reveals elevated interferon responses and TNF-α signaling via NFkB in monocytes in children with uncomplicated malaria"

### Supplementary_Figure_1

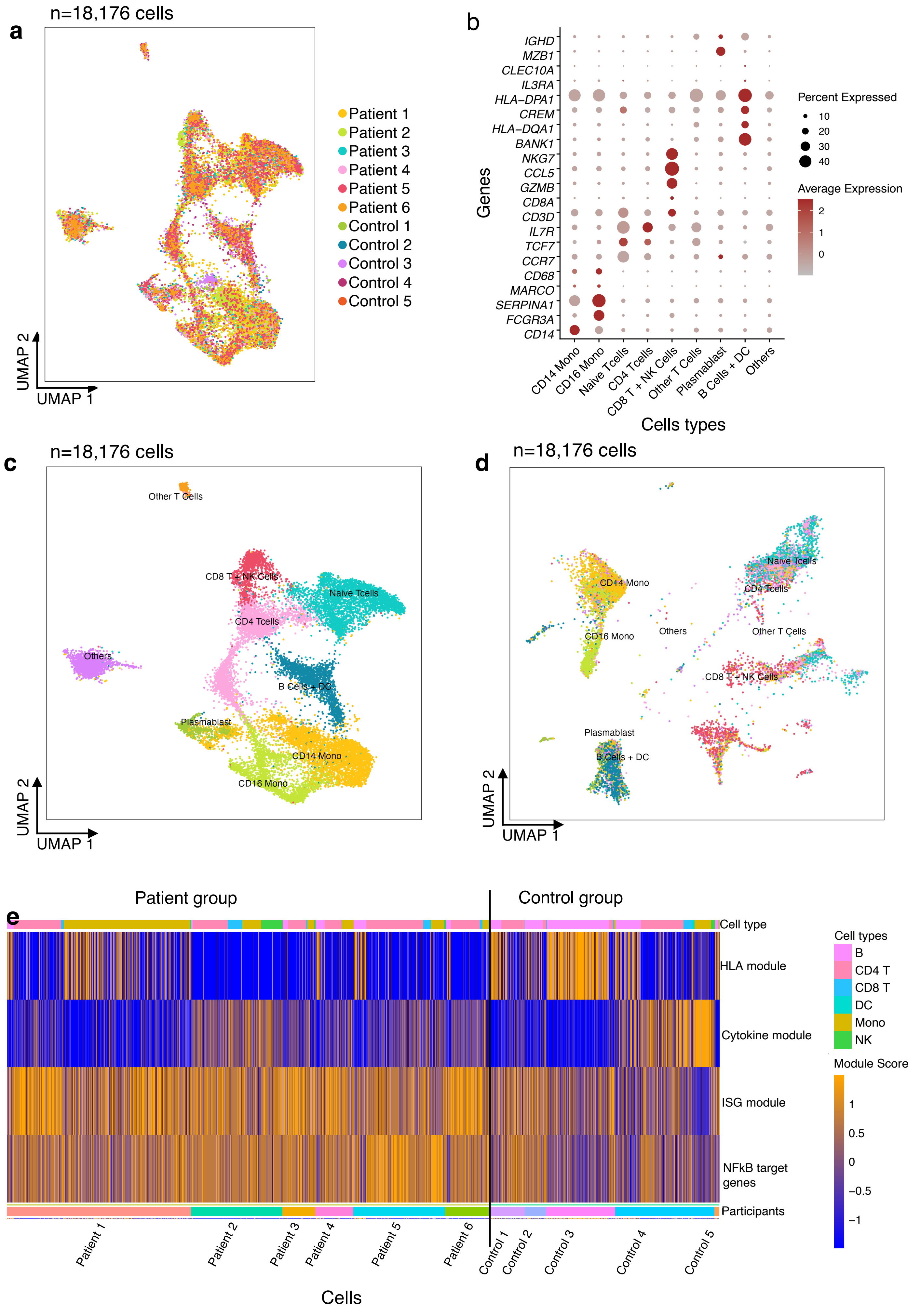
